# Prevalence and associated factors of gender-based violence for female: Evidence from school students in Nepal- a cross sectional study

**DOI:** 10.1101/2024.05.14.24307359

**Authors:** Laxmi Gautam, Manisha Sha, Durga Khadka Mishra, Padam Kanta Dahal, Sujan Gautam

**Affiliations:** Department of Public Health, Manmohan Memorial Institute of Health Sciences, Kathmandu, Nepal; Madan Bhandari Academy of Health Sciences, Hetauda, Nepal; School of Health, Medical and Applied Sciences, Central Queensland University, Sydney Campus, NSW, Australia

**Keywords:** Gender-based violence, secondary school female students, sexual violence, physical violence, emotional violence

## Abstract

**Background:** Gender-based violence (GBV) is a major global public health challenge in 21^st^ century that remains a serious impact on women’s health and well-being. Therefore, this study aimed to access the prevalence and factors associated with GBV among secondary school female students in Sarlahi district of Nepal.

**Methods:** Using a cross-sectional study, we collected data from 225 secondary school female students in Sarlahi district of Nepal. Data was collected by using a semi-structured, self-administered questionnaire. Probability proportionate and simple random sampling techniques were used for sampling. The association were explored by using chi-square test and binary logistic regression where a p value <0.05 was considered statistically significant.

**Results:** The prevalence of GBV among the students during their lifetime was 45.33% followed by physical (16.89%), sexual (30.22%) and psychological (39.56%) violence respectively. The prevalence of physical violence from family members was 97.36%, followed by emotional violence (41.57%). Further, the prevalence of sexual violence from the non-family members was 91.17%. Type of family had a significant association with lifetime experience of GBV (p=0.003). Gender based discrimination in the family had a significant association with lifetime (p=0.001) as well as last 12 months (p=0.001) GBV experience. Experience of witnessing physical violence as a child was within last 12 months was associated with GBV (p=0.03).

**Conclusion:** GBV has been prevalent among the high school students, with limited level of awareness in that issue. This warrants the urgent need to establish preventive and responsive control measure within schools and communities to address the GBV effectively.

## Introduction

Gender-based violence (GBV) remains one of the most serious social, legal and health challenges for the 21^st^ century. It is a major public health problem and human right concern throughout the world which has a serious impact on women’s health and well-being.^(1)^ GBV or violence against women and girls (VAWG), is a global pandemic that affects 1 in 3 women in their lifetime and 35% of women worldwide have experienced either physical and/or sexual intimate partner violence or non-partner sexual violence. Further, world health organization (WHO) estimated that 1 in 5 women between 15-24 years could subject to GBV from their intimate partners while 40.2% of all women experience physical and/or sexual intimate partner or non-partner violence in their lifetime in southeast Asia.^(2)^ (REF) Studies in India^(3)^, Pakistan^(4)^ and Maldives^(5)^ revealed that 79.6%, 38.40% and 34.6% respectively.

In Nepal, the prevalence of GBV is rapidly increased over few decades which has created a serious impact in the health and well-being of females. Evidence shows that over 150 females were killed as a result of GBV in 2017, highlighted the major perpetrator was family member or relatives.^(6)^ Further, another study conducted among community people in Kathmandu, Lalitpur, Bhaktapur, and Nuwakot districts in 2018 showed 11.3% respondent experience GBV in past 12 months of survey.^(7)^ Nepal Demographic Health Survey in 2022 showed that 23% of women in aged 15–49 have experienced physical violence since age of 15 and 8% which is a percentage higher compared to 2016 estimates.^(8),(9)^

GBV occurs as a result of the normative role expectations associated with each gender and unequal power relationships within the context of a specific society. Domestic violence, marital rape, dowry-related violence, child marriage, polygamy, female infanticide, witchcraft accusations, chhaupadi, trafficking of women and girls for sexual exploitation are common types of GBV in Nepal.^(10)^ Furthermore, patriarchal attitudes and deep-rooted stereotypes that discriminate against women remain entrenched in the social, cultural, religious, socio-economic and political institutions. However, studies are lacking to estimate the prevalence of gender-based violence on the school level in low and middle-income countries like Nepal and insufficient for designing the responsive control intervention to fight against the problem. Therefore, this study aimed to assess the prevalence and factor associated with gender-based violence among secondary school’s female students.

## Materials and Methods

### Study design and population

A cross-sectional study was conducted from October 2020 to January 2021 among female students of grade 9 to 12 in Brahmpuri Rural Municipality of Sarlahi district, Nepal. A sample of 225 participant was estimated using Cochran’s formula with reference of prevalence of GBV (Prevalence =48%) from a study conducted by Government of Nepal^(11)^ at 5% allowable error and 10% non-response rate. Two schools were selected using simple random sampling and probability proportionate allocation (PPA) was used to select the study population. Pretested semi-structured questionnaire were provided for self-administration. Information about socio-demographic status, violence related exposure and experience along with prevalence of GBV in lifetime and last 12 months preceding survey were collected. In addition, the study was adhered to the Strengthening the Reporting of Observational Studies in Epidemiology (STROBE) guidelines for reporting observational studies

### Outcome measures

Gender-based violence was the major outcome of this study including the prevalence of physical, Sexual and Psychological violence. In this study physical violence includes if the respondents reported to have experienced slapping, punching, kicking/dragging, beating/hitting with any object, cutting/ biting, shaking, shoving, pushing, throwing, and burning/chocking against them. Similarly for sexual violence it was considered if the respondent had experienced unwanted or non-consensual sexual act through force or threat against them. In case of psychological violence if there was systemic destruction of women’s or girl’s self-esteem and/or sense of safety ten it was considered as violence against them which included humiliation, being made to feel unwanted, forced isolation from family or friends, threats to harm the individual or someone they care about, repeated yelling or degradation, inducing fear through intimidating words or gestures, controlling behavior, and the destruction of possessions. To measure these experiences different questions were developed based on the reference provided by WHO, UNICEF and UN women and were modified to make applicable in local context of the respondents. The Prevalence of GBV was measured if the respondents had experiences any type of physical, sexual, psychological violence that targets them.

### Statistical analyses

The frequency, percentage, mean and standard deviation were reported for each variable. The association between dependent and independent variables were determined by chi-square test and binary logistic regression. Bivariate and multivariate analyses were conducted and the association were tested for different independent variables with the experience of violence in lifetime and last 12 months of the survey. The probability value less than 0.05 was considered to be significant. Microsoft excel was used for data cleaning and coding and statistical package of social science (SPSS) version 16 was used for statistical analyses.

### Ethical Statement

Ethical approval was received from institutional review committee (IRC) of Manmohan Memorial Institute of Health Sciences (MMIHS-IRC 77/13). Study was conducted from October 2020 to January 2021. Further, permission was taken from each school and municipality before the information collection. Written informed consent was consent was taken from participants after explaining purpose of the study prior to the data collection. In case of participants below the age of 18 years, permission was taken from school principle, respective class teachers and then assent was taken from them. Participants were informed about privacy and confidentiality of information as well as the right of withdrawal and right of refusal. All other ethical guidelines were followed throughout the process of this study.

## Results

The mean age of respondents was 17.17±2.48 years with more than two third of them were from age group 15-20 years. Father and grandfather were the major decision makers of their family. Alcohol and drugs abusers in the family were reported by more than one third respondents. Most of the respondents need permission from family to go to health institution while more than a quarter (28.4%) faced discrimination in comparison to their male counterparts. Almost 30% responded had experienced witnessed GBV as a child, followed by sexual (10.7%) and physical violence (8.9%) **(Table 1)**.

**Table 1:** Socio-demographic information and Bivariate analysis GBV ever among the respondents.

| Variables | GBV ever |  | Total | P value |
| --- | --- | --- | --- | --- |
|  | Yes (%) | No (%) |  |  |
| Age in years |  |  |  |  |
| <15 | 25 (60.98) | 16 (39.02) | 41 (18.2) | 0.001 |
| 15-20 | 90 (59.2) | 62 (40.8) | 152 (67.6) |  |
| >20 | 8 (25.0) | 24 (75.0) | 32 (14.2) |  |
| Religion |  |  |  |  |
| Hindu | 75 (39.7) | 114 (60.3) | 189 (84.0) | <0.001 |
| Other than Hindu | 27 (75.0) | 9 (25.0) | 36 (16.0) |  |
| Type of Family |  |  |  |  |
| Nuclear family | 46 (36.5) | 80 (63.5) | 126 (56.0) | 0.003 |
| Joint family and extended | 56 (56.6) | 43 (43.4) | 99 (44.0) |  |
| Living status of Parents |  |  |  | <0.001 |
| Together | 65 (35.5) | 118 (64.5) | 183 (81.3) |  |
| Separately | 37 (88.1) | 5 (11.9) | 42 (18.7) |  |
| Educational status of father (n=222) |  |  |  |  |
| Higher Education (bachelors and above) | 1 (10.0) | 9 (90.0) | 10 (4.5) | <0.001 |
| Secondary Education (9-12) | 14 (18.7) | 61 (81.3) | 75 (33.8) |  |
| Primary Education (1-8) | 47(55.3) | 38 (44.7) | 85 (38.3) |  |
| Literate (able to read and write) | 17 (60.7) | 11 (39.3) | 28 (12.6) |  |
| Illiterate (unable to read or write) | 20 (83.3) | 4 (16.7) | 24 (10.8) |  |
| Educational status of Mother (n=219) |  |  |  | <0.001 |
| Secondary Education (9-12) | 5 (31.3) | 11 (68.8) | 16 (7.3) |  |
| Primary Education (1-8) | 14 (18.7) | 61 (81.3) | 75 (34.2) |  |
| Literate (able to read or write) | 28 (48.3) | 30 (51.7) | 58 (26.5) |  |
| Illiterate (unable to read or write) | 49 (70.0) | 21 (30.3) | 70 (32.0) |  |
| Decision maker of the family |  |  |  | 0.002 |
| Father/Grandfather | 83 (41.3) | 118 (58.7) | 201 (89.3) |  |
| Mother/Grandmother | 11 (73.3) | 4 (26.7) | 15 (6.7) |  |
| Others | 8 (88.9) | 1 (11.1) | 9 (4.0) |  |
| Alcohol or drugs abuser in the family |  |  |  |  |
| No | 36 (26.1) | 102 (73.9) | 138 (61.3) | <0.001 |
| Yes | 66 (75.9) | 21 (24.1) | 87 (38.7) |  |
| Need permission from family members to visit health center or hospital |  |  |  |  |
| No | 0 (0) | 4 (100) | 4 (1.8) | 0.066 |
| Yes | 102 (46.2) | 119 (53.8) | 221 (98.2) |  |
| Discriminate between men and women in family or treats them differently |  |  |  |  |
| No | 48 (29.8) | 113 (70.2) | 161 (71.6) | <0.001 |
| Yes | 54 (84.4) | 10 (15.6) | 64 (28.4) |  |
| Experience of Witnessing GBV as a child (n=218) |  |  |  |  |
| No | 51 (32.9) | 104 (67.1) | 155 (71.1) | <0.001 |
| Yes | 47 (74.6) | 16 (25.4) | 63 (28.9) |  |
| Experience of witnessing physical violence as a child |  |  |  |  |
| No | 84 (40.8) | 122 (59.2) | 205 (91.1) | <0.001 |
| Yes | 18 (94.7) | 1 (5.3) | 20 (8.9) |  |
| Experience of witnessing sexual violence as a child |  |  |  |  |
| No | 79 (39.3) | 122 (60.7) | 201 (89.3) | <0.001 |
| Yes | 23 (95.8) | 1 (4.2) | 24 (10.7) |  |

Regarding the GBV with in last 12 months preceding the survey, age and prevalence of violence had a direct association. Similarly, GBV was experienced more in joint family, and was significantly associated with those whose parents were separated, educational status of parents, existence of alcohol or drugs abuser in the family, discriminate between men and women in family, and experience of witnessing GBV as a child in last 12 months before the survey **(Table 2)**.

**Table 2:**
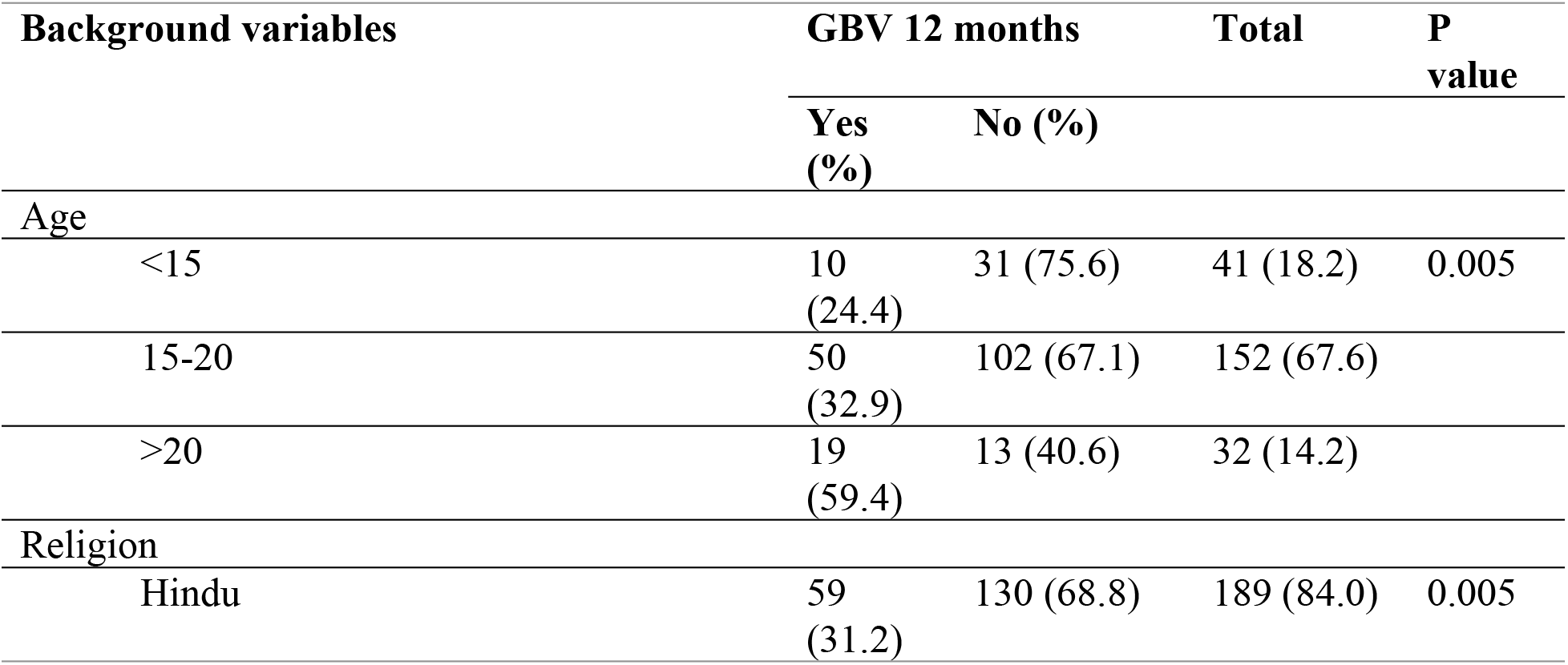
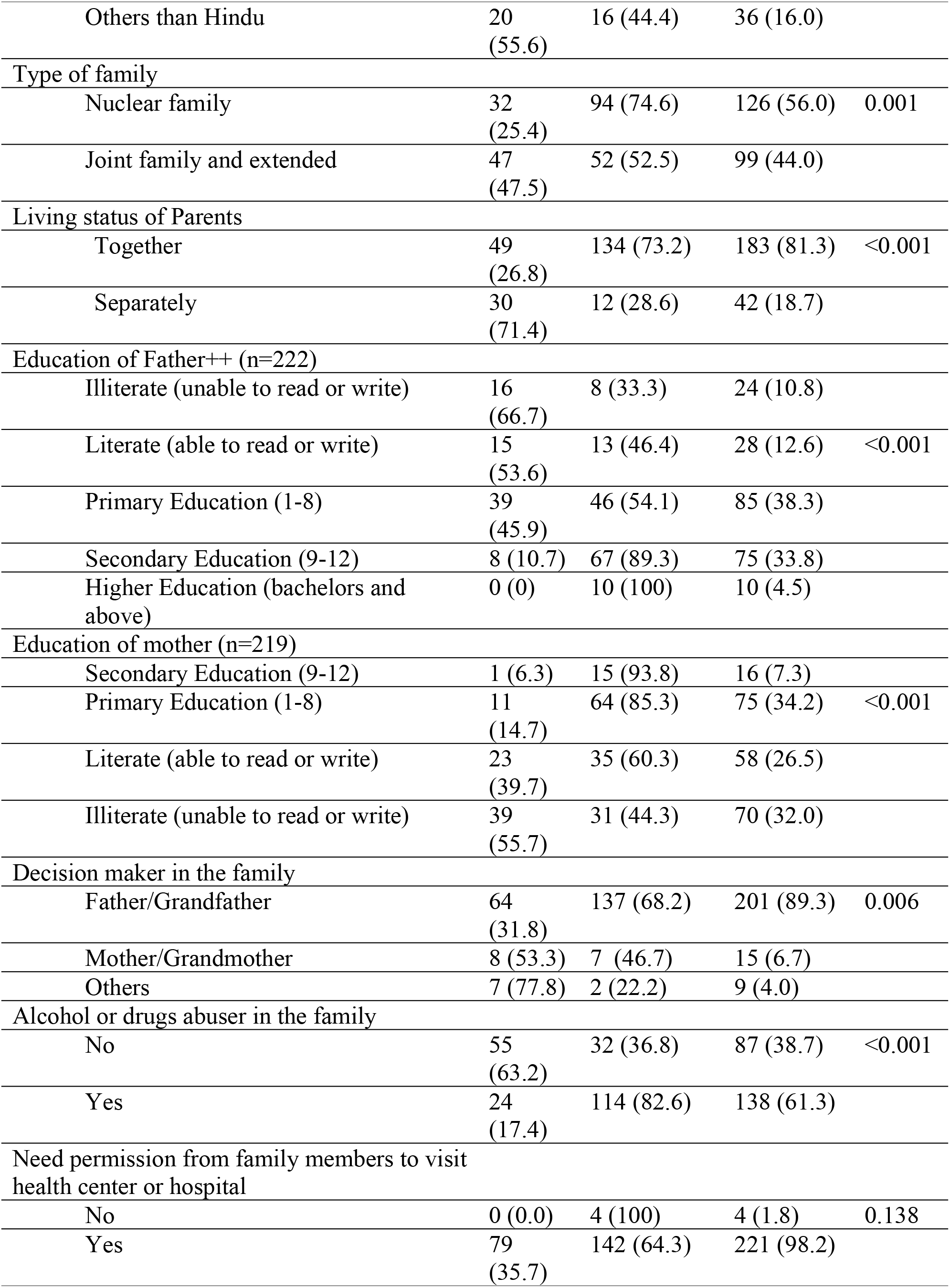
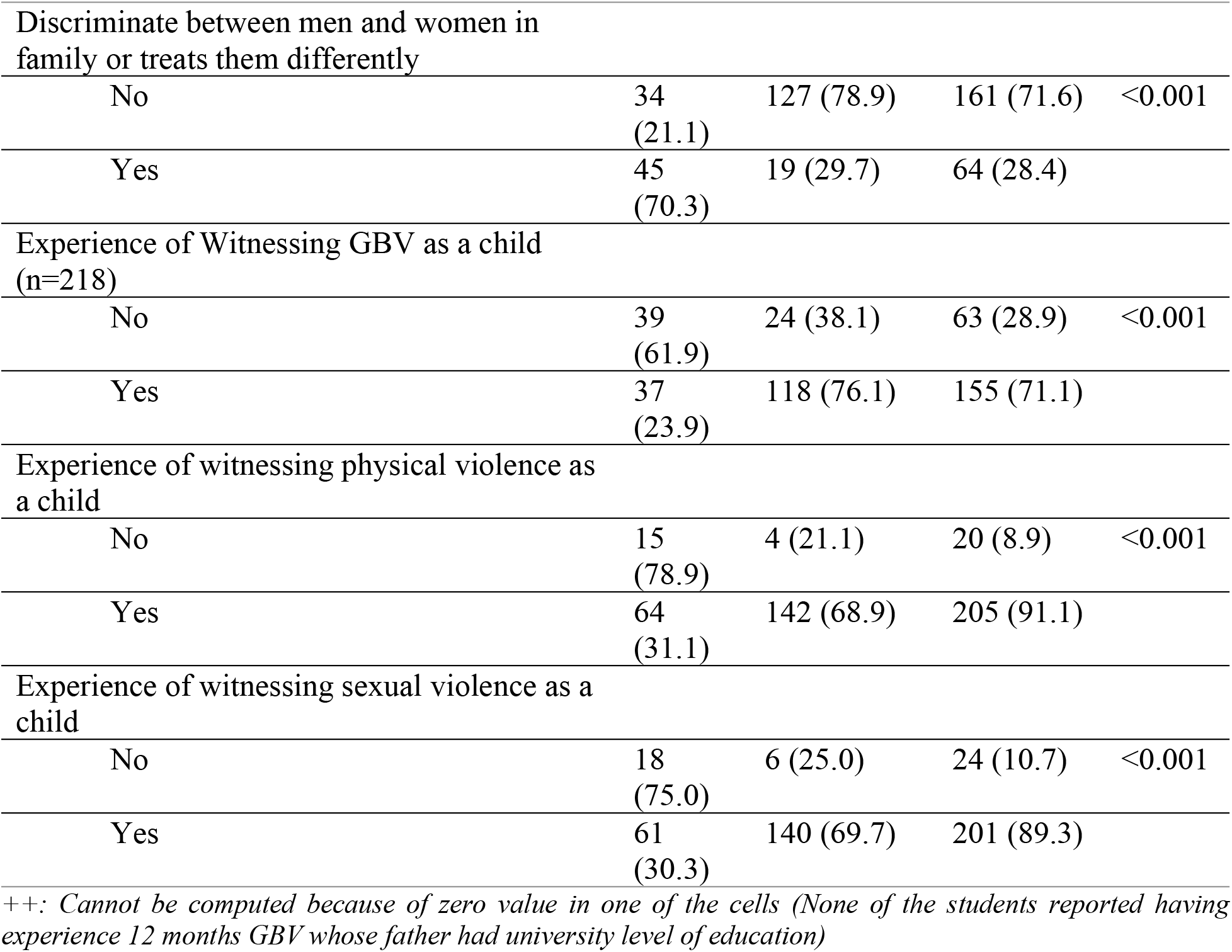
Bivariate analysis GBV within last 12 months preceding the survey.

GBV in lifetime was 4.68 (1.69-12.95) times more likely to happen among respondents aged more than 20 compared to < 15 years age group. Similarly, there was 4.56 (2.03-10.23) times more likely to face GBV by non-Hindu by religion in comparison to Hindu respondents. Respondents whose parents were not living together were likely to face GBV 13.43 (5.03-35.85) times than those whose parents were living together. The higher the education of parents the lesser the chances of facing GBV. The alcohol or drugs abuser in family was also contributed to increase the likelihood of GBV by 8.91 (4.78-16.57) times and discrimination on family among men and women by 12.71 (5.97-27.03) times. Regarding the childhood experience of witnessing violence, those who had witness GBV were 5.99 (3.01-11.57) times more at odd of facing GBV later. In case of GBV 12 months preceding survey, odd of facing GBV was increasing along with the age, was more among religion other than Hindu and more among joint family compared to nuclear family. Those respondents whose parents were not together were more at risk of facing GBV (6.84 (3.24-14.40)) than those whose parents were living together. Alcohol or drugs abuser in family had increased the odd of GBV by 8.16 (4.39-15.16) times similarly those respondents who faced discrimination in family were 8.08 (4.58-17.05) times more likely to face GBV compared to those who didn’t face discrimination within 12 months prior to the survey. Type of family had significant association with lifetime experience of GBV (p=0.02) while living status of parents was associated with 12 months experience of GBV (p=0.02) and decision maker in family was associated with both. Discriminate between men and women in family had significant association with lifetime and 12 months GBV experience (p=0.01 and p=0.001) **(Table 3)**.

**Table 3:**
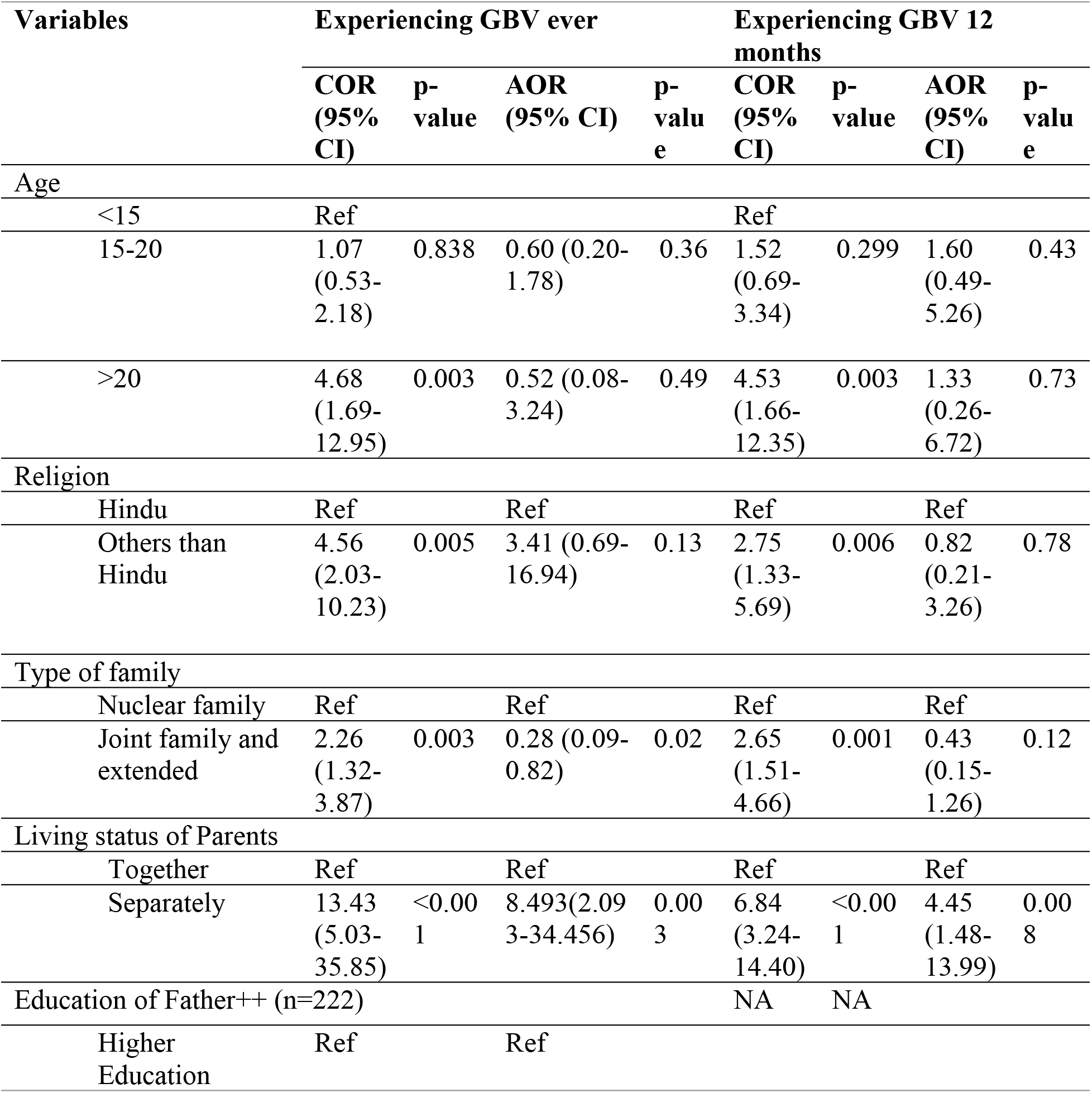
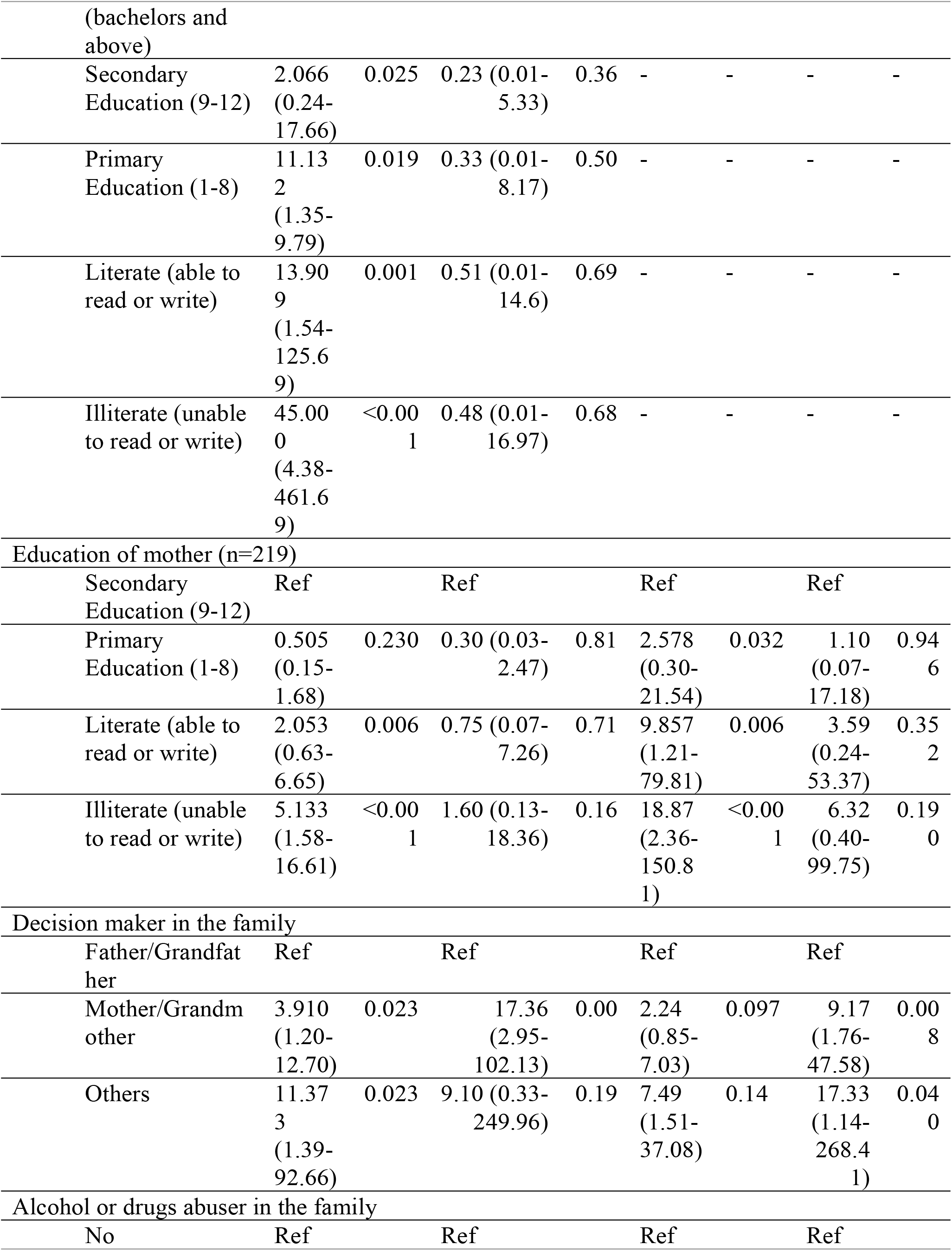
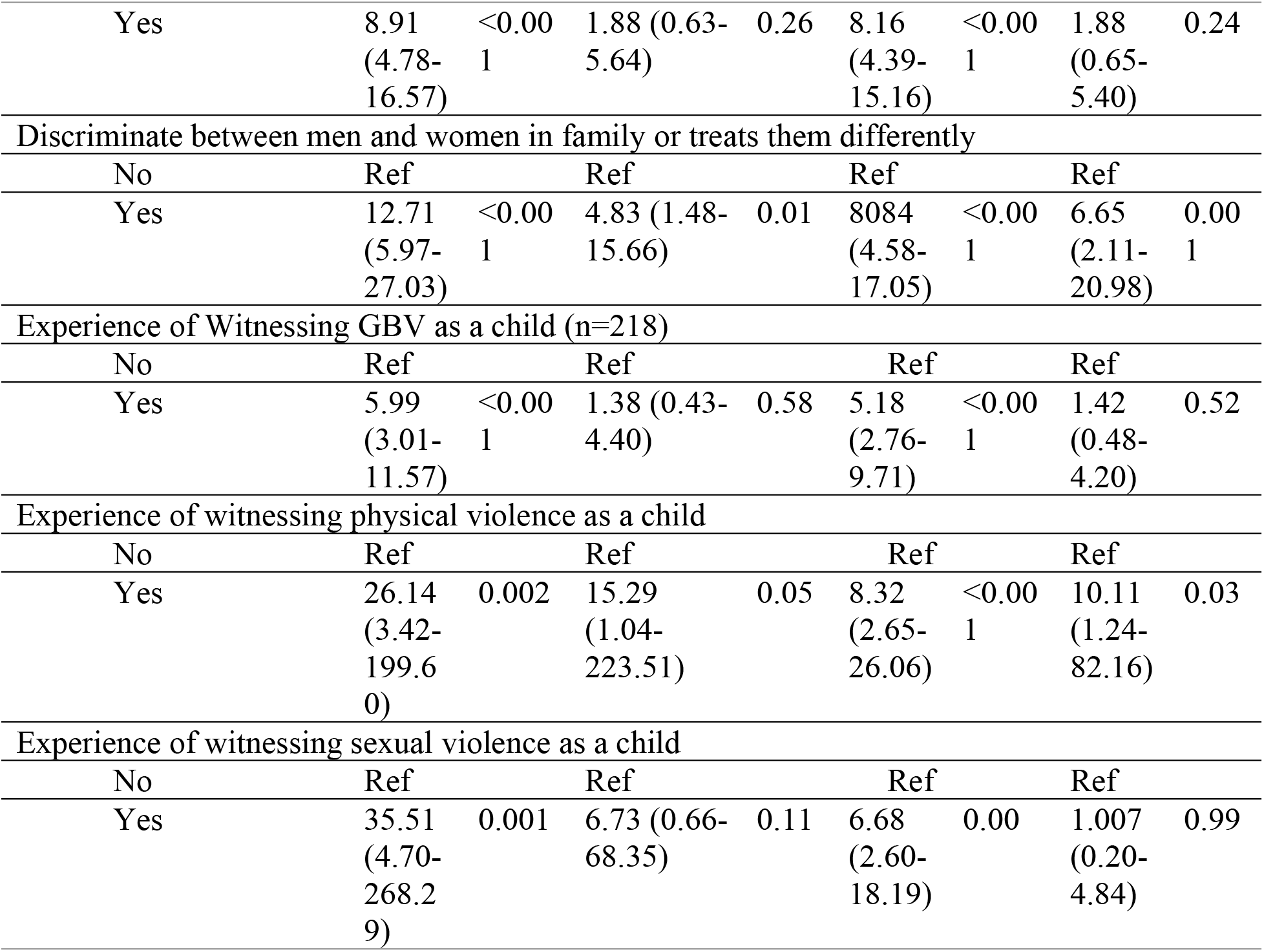
Multivariate GBV ever and within last 12 Months.

| Variables | Experiencing GBV ever |  |  |  | Experiencing GBV 12 months |  |  |  |
| --- | --- | --- | --- | --- | --- | --- | --- | --- |
|  | COR (95% CI) | p-value | AOR (95% CI) | p-value | COR (95% CI) | p-value | AOR (95% CI) | p-value |
| **Age** |  |  |  |  |  |  |  |  |
| <15 | Ref |  |  |  | Ref |  |  |  |
| 15-20 | 1.07 (0.53-2.18) | 0.838 | 0.60 (0.20-1.78) | 0.36 | 1.52 (0.69-3.34) | 0.299 | 1.60 (0.49-5.26) | 0.43 |
| >20 | 4.68 (1.69-12.95) | 0.003 | 0.52 (0.08-3.24) | 0.49 | 4.53 (1.66-12.35) | 0.003 | 1.33 (0.26-6.72) | 0.73 |
| **Religion** |  |  |  |  |  |  |  |  |
| Hindu | Ref |  | Ref |  | Ref |  | Ref |  |
| Others than Hindu | 4.56 (2.03-10.23) | 0.005 | 3.41 (0.69-16.94) | 0.13 | 2.75 (1.33-5.69) | 0.006 | 0.82 (0.21-3.26) | 0.78 |
| **Type of family** |  |  |  |  |  |  |  |  |
| Nuclear family | Ref |  | Ref |  | Ref |  | Ref |  |
| Joint family and extended | 2.26 (1.32-3.87) | 0.003 | 0.28 (0.09-0.82) | 0.02 | 2.65 (1.51-4.66) | 0.001 | 0.43 (0.15-1.26) | 0.12 |
| **Living status of Parents** |  |  |  |  |  |  |  |  |
| Together | Ref |  | Ref |  | Ref |  | Ref |  |
| Separately | 13.43 (5.03-35.85) | <0.001 | 8.493 (2.09-34.456) | 0.003 | 6.84 (3.24-14.40) | <0.001 | 4.45 (1.48-13.99) | 0.008 |
| **Education of Father++ (n=222)** |  |  |  |  | NA | NA |  |  |
| Higher Education | Ref |  | Ref |  |  |  |  |  |

|  |  |  |  |  |  |  |  |  |  |
| --- | --- | --- | --- | --- | --- | --- | --- | --- | --- |
| (bachelors and above) |  |  |  |  |  |  |  |  |  |
| Secondary Education (9-12) | 2.066<br>(0.24-17.66) | 0.025 | 0.23 (0.01-5.33) | 0.36 | - | - | - | - | - |
| Primary Education (1-8) | 11.13<br>2<br>(1.35-9.79) | 0.019 | 0.33 (0.01-8.17) | 0.50 | - | - | - | - | - |
| Literate (able to read or write) | 13.90<br>9<br>(1.54-125.69) | 0.001 | 0.51 (0.01-14.6) | 0.69 | - | - | - | - | - |
| Illiterate (unable to read or write) | 45.00<br>0<br>(4.38-461.69) | <0.001 | 0.48 (0.01-16.97) | 0.68 | - | - | - | - | - |
| Education of mother (n=219) |  |  |  |  |  |  |  |  |  |
| Secondary Education (9-12) | Ref |  | Ref |  | Ref |  | Ref |  |  |
| Primary Education (1-8) | 0.505<br>(0.15-1.68) | 0.230 | 0.30 (0.03-2.47) | 0.81 | 2.578<br>(0.30-21.54) | 0.032 | 1.10<br>(0.07-17.18) | 0.94<br>6 |  |
| Literate (able to read or write) | 2.053<br>(0.63-6.65) | 0.006 | 0.75 (0.07-7.26) | 0.71 | 9.857<br>(1.21-79.81) | 0.006 | 3.59<br>(0.24-53.37) | 0.35<br>2 |  |
| Illiterate (unable to read or write) | 5.133<br>(1.58-16.61) | <0.001 | 1.60 (0.13-18.36) | 0.16 | 18.87<br>(2.36-150.81) | <0.001 | 6.32<br>(0.40-99.75) | 0.19<br>0 |  |
| Decision maker in the family |  |  |  |  |  |  |  |  |  |
| Father/Grandfather | Ref |  | Ref |  | Ref |  | Ref |  |  |
| Mother/Grandmother | 3.910<br>(1.20-12.70) | 0.023 | 17.36<br>(2.95-102.13) | 0.00 | 2.24<br>(0.85-7.03) | 0.097 | 9.17<br>(1.76-47.58) | 0.00<br>8 |  |
| Others | 11.37<br>3<br>(1.39-92.66) | 0.023 | 9.10 (0.33-249.96) | 0.19 | 7.49<br>(1.51-37.08) | 0.14 | 17.33<br>(1.14-268.41) | 0.04<br>0 |  |
| Alcohol or drugs abuser in the family |  |  |  |  |  |  |  |  |  |
| No | Ref |  | Ref |  | Ref |  | Ref |  |  |
| Yes | 8.91<br>(4.78-16.57) | <0.00<br>1 | 1.88 (0.63-5.64) | 0.26 | 8.16<br>(4.39-15.16) | <0.00<br>1 | 1.88<br>(0.65-5.40) | 0.24 |
| Discriminate between men and women in family or treats them differently |  |  |  |  |  |  |  |  |
| No | Ref |  | Ref |  | Ref |  | Ref |  |
| Yes | 12.71<br>(5.97-27.03) | <0.00<br>1 | 4.83 (1.48-15.66) | 0.01 | 8.084<br>(4.58-17.05) | <0.00<br>1 | 6.65<br>(2.11-20.98) | 0.00<br>1 |
| Experience of Witnessing GBV as a child (n=218) |  |  |  |  |  |  |  |  |
| No | Ref |  | Ref |  | Ref |  | Ref |  |
| Yes | 5.99<br>(3.01-11.57) | <0.00<br>1 | 1.38 (0.43-4.40) | 0.58 | 5.18<br>(2.76-9.71) | <0.00<br>1 | 1.42<br>(0.48-4.20) | 0.52 |
| Experience of witnessing physical violence as a child |  |  |  |  |  |  |  |  |
| No | Ref |  | Ref |  | Ref |  | Ref |  |
| Yes | 26.14<br>(3.42-199.6<br>0) | 0.002 | 15.29<br>(1.04-223.51) | 0.05 | 8.32<br>(2.65-26.06) | <0.00<br>1 | 10.11<br>(1.24-82.16) | 0.03 |
| Experience of witnessing sexual violence as a child |  |  |  |  |  |  |  |  |
| No | Ref |  | Ref |  | Ref |  | Ref |  |
| Yes | 35.51<br>(4.70-268.2<br>9) | 0.001 | 6.73 (0.66-68.35) | 0.11 | 6.68<br>(2.60-18.19) | 0.00 | 1.007<br>(0.20-4.84) | 0.99 |

Almost half (45.3%) of the respondents had experienced GBV in lifetime followed by last 12 months (35.1%) preceding survey. The lifetime physical, sexual and psychological violence among the respondents was 16.89%, 30.22% and 39.56% respectively. Regarding the perpetrators of GBV, physical violence was mainly from family members (97%) while sexual violence was mainly from non-family members (91%). Less than half (42.15%) of the respondents had reported to someone after violence because 38.98% had no idea that was crime 20.33% did not report due to fear **(Table 4)**.

**Table 4:** Prevalence of different types of GBV, perpetrators and reporting status after violence.

| Variables | Frequency | Percentage (%) |
| --- | --- | --- |
| Ever Experienced GBV in lifetime |  |  |
| Yes | 102 | 45.3 |
| No | 123 | 54.7 |
| Experienced GBV in last 12 months preceding study |  |  |
| Yes | 79 | 35.1 |
| No | 146 | 64.9 |
| Physical violence |  |  |
| Yes | 38 | 16.89 |
| No | 187 | 83.11 |
| Physical Violence in last 12 months<br>(n=38) |  |  |
| Yes | 14 | 36.8 |
| No | 24 | 63.2 |
| Sexual violence |  |  |
| Yes | 68 | 30.22 |
| No | 157 | 69.78 |
| Sexual Violence In last 12 months<br>(n=49) |  |  |
| Yes | 39 | 57 |
| No | 29 | 43 |
| Emotional violence |  |  |
| Yes | 90 | 40 |
| No | 135 | 60 |
| Emotional Violence in last 12 months<br>(n=90) |  |  |
| Yes | 60 | 66.67 |
| No | 30 | 33.33 |
| Perpetrators |  |  |
| Physical violence |  |  |
| Family member | 37 | 97 |
| Non-Family members | 1 | 3 |
| Sexual violence |  |  |
| Family member | 6 | 9 |
| Non-Family members | 62 | 91 |
| Emotional violence |  |  |
| Family member | 37 | 42 |
| Non-Family members | 53 | 58 |
| Reporting after Violence (n=102) |  |  |
| Yes | 43 | 42.15 |
| No | 59 | 57.84 |
| Reasons for not reporting violence (n=59) |  |  |
| No idea that was crime | 23 | 38.98 |
| Scared to report | 12 | 20.33 |
| Because of family prestige | 11 | 18.64 |
| Embarrassment | 7 | 11.86 |
| Accept violence as normal | 6 | 10.16 |

## Discussion

To our knowledge, this is the first study to explore the prevalence and factor of GBV among the secondary school female students in Nepal. This study identified that the risk of GBV including physical, mental and sexual violence increasing among the secondary school girls where the joint family status, poor education status of parents, single parents and existence of alcohol or drugs abusers in the family. Further, GBV among the teenage girls is highly prevalent in both in lifetime and 12 months prior to the survey.

This study identified the higher risk of GBV among the girls who are studying in higher secondary level, which is supported by the gender gap index 0.659 in 2023.^(12)^ In line to our current study findings, previous studies conducted in Debre Markos Town^(13)^, Nepal^(11)^ and Ethiopia^(14)^ showed that the prevalence of GBV among girls was 47%, 48% and 47.2% respectively. However, our estimates are relatively lower than the study conducted in Southeast Ethiopia (68.2%) and Northwest Ethiopia (67.7%.4).^(15), (16)^ These variations might be due to the difference in status of women and gender gap.

Alike to our study, previous studies conducted in Ethiopia^(17),(14)^ and South Africa^(18)^ showed that experience of physical violence among the students were 36.2%, 37.99% and 36.3% respectively. Further, Nepal demographic health survey (NDHS) 2022 also prevailed that 23% of women aged 15 to 49 have experienced physical violence since age of 15, including 11% experienced physical violence.^(8)^ This survey also highlighted that 37% women of reproductive age experienced physical violence in Madhesh Province where this study was conducted. In addition, physical violence on both lifetime and 12 months prior to our study are comparable to the study conducted in Kenya which were 18.0 and 10.7% respectively.^(19)^ The risk of sexual violence is similar to the previous study conducted in Ethiopia (27.99%).^(14)^ In contrast, study in South Africa showed 46% prevalence of sexual violence.^(18)^ The higher prevalence of sexual violence in these studies could be prevailed because of the different study population who were university students. University students are more mature than school students to start intimate relationship and have more risk of sexual violence because of initiation of intimate relationship.^(20)^ Furthermore, NDHS 2022 showed that 8% of women aged 15 to 49 have ever experienced sexual violence, and 4% experienced sexual violence in the 12 months preceding the survey which was 11% in Madesh province.^(8)^ The experience of emotional violence in our current study is lower by 50% compared to the previous studies in Ethiopia and South Africa.^(17),(18)^ However, NDHS reported that the proportion of women who reported being forced to get married is highest in Madhesh Province (20%), not cared during illness (11.5%), threatened for divorce (10%) were reported by women of reproductive age which might be the drivers for the emotional violence.^(8)^

Present study found that those respondents who had discriminate between men and women in family were more at risk of GBV which was supported by a study conducted in Nepal which showed that a power play between men and women reinforce inequality and increases the likelihood of violence for women.^(21)^ Further, a study conducted in Sub Saharan Africa showed that having an illiterate mother [OR 2.13] was associated with violence which is similar to the current study.^(4)^ Similarly, respondents whose father had university level of education did not face GBV with in last 12 months and the risk of GBV was inversely proportional with the education of father. This finding was supported by a study in Sub-Sharan Africa; having a father who had completed primary or lower school was three or four times more likely to experience [OR 3.06 and 4.69] sexual violence respectively then compared to father with higher studies.^(22)^ This might be due to mothers with low educational status may be less aware about GBV and unable to teach their daughter about this issue and protect them while they accept violence as normal process themselves. There are not much study conducted to examine the childhood exposure to violence and its factors in Nepalese context. However, one of the largest investigations ever conducted on childhood trauma the adverse childhood experiences (ACE) study in the US showed that experience of childhood trauma, separation of parents, substance abusing, experience of domestic violence are the risk factors of GBV among children which in line to the findings of present study.^(23)^ Similarly, students who witnessed physical violence during childhood was (OR: 15.29, CI: 1.04-223.51) more likely to feel violence in their life time and also have significant association with GBV within the last 12 months before the survey (p=0.03). In case of experience of witnessing childhood sexual violence, the risk of GBV was 6.73 (0.66-68.35) times higher than those who did not witness such violence in childhood.^(23)^ A systematic review and meta-analysis in education institution of Sub-Sharan Africa shows in three studies, researchers found witnessing parental violence [OR 2.40, 2.20, and 1.54] was associated with an increased risk of GBV.^(22)^

This study showed that the childhood exposure to witnessing violence was 1.38 times more likely experience GBV in comparison to those who were not exposed to GBV. This finding was supported by the study conducted by Temesgen in Ethiopia which showed that female students who had witness parental violence were 1.92 times more likely to experience GBV as compared to those who didn’t witness any paternal violence.^(24)^ Substance abuse seems as a risk factor of GBV in this study with risk of 1.88 times among the respondents from family with abuser which was supported by a study conducted in Ethiopia and Sub-Saharan African.^(22), (23)^ This may be due to alcohol influencing decision making, and person lose their control among themselves which may lead to GBV. Our study showed that the offenders of physical and sexual violence were mostly from family members which was in line with the finding previous study which showed that family members as offenders of physical violence was 50.2% which was 21.2% in case of sexual violence.^(25)^ This might be due to the cultural and traditional belief among family member that highlighted the importance of existing the strong policy implementation^(26)^ and provide sufficient protection from multiple and intersecting forms of discrimination for women and girls. Even though the information collector was from local community some respondents may have underreported their experiences because the issue was sensitive to discuss openly. In addition the study was conducted in some school among female students only which was not able to address the issues of children out of school as well as knowledge and perception of male students. So the findings may not represent the situation of overall children which shows the need of larger level study by including all mentioned participants.

The Constitution of Nepal 2072, gives right to no woman shall be subjected to physical, mental, sexual, psychological or other form of violence or exploitation. ^(27)^ Children are future adult and it is necessary to prevent them from any type of violence because such experiences can have intergenerational effects. After school students will start intimate relationship and will have their own family in future which can put them in more risk of violence because major perpetrators of violence are intimate partners. So school students are required to make aware about the GBV, its effect in their life and preventive measures because majority of them were not aware that they are the victim of GBV and it should be reported.

## Conclusions

In conclusion, GBV including physical, sexual and emotional violence are prevalent among the female secondary students. The drivers of physical violence were mainly from family members while sexual violence was from non-family members. Joint or extended types of family, discrimination between the gender within the family, living and education status of parents, and experience of witnessing violence as a child were main factors associated to the GBV. This warrants the urgent needs of awareness to the community regarding the female rights for their self-protection, protection from exposure to GBV since childhood and education on the gender equality within the family. Further, strong policy implementation to protect the girls from the violence and proper reporting mechanism through school and other appropriate channel is recommended.

## Data Availability

All the data generated in the study are included in the manuscript.

## Acknowledgements

Special thanks to Education officer Mr. Jay Prakash Shah and other teachers who supported me during data collection as well as all the schools for their support. The support from all the members of faculty of public health, Manmohan Memorial Institute of Health Sciences is also appreciative. Above all, cordial thanks to all the respondents for their time without whom this study would not have been possible.

